# A holistic approach for suppression of COVID-19 spread in workplaces and universities

**DOI:** 10.1101/2020.12.03.20243626

**Authors:** Sarah F. Poole, Jessica Gronsbell, Dale Winter, Stefanie Nickels, Roie Levy, Bin Fu, Maximilien Burq, Sohrab Saeb, Matthew D. Edwards, Michael K. Behr, Vignesh Kumaresan, Alexander R. Macalalad, Sneh Shah, Michelle Prevost, Nigel Snoad, Michael P. Brenner, Lance J. Myers, Paul Varghese, Robert M. Califf, Vindell Washington, Vivian S. Lee, Menachem Fromer

## Abstract

As society has moved past the initial phase of the COVID-19 crisis that relied on broad-spectrum shutdowns as a stopgap method, industries and institutions have faced the daunting question of how to return to a stabilized state of activities and more fully reopen the economy. A core problem is how to return people to their workplaces and educational institutions in a manner that is safe, ethical, grounded in science, and takes into account the unique factors and needs of each organization and community. In this paper, we introduce an epidemiological model (the “Community-Workplace” model) that accounts for SARS-CoV-2 transmission within the workplace, within the surrounding community, and between them. We use this multi-group deterministic compartmental model to consider various testing strategies that, together with symptom screening, exposure tracking, and nonpharmaceutical interventions (NPI) such as mask wearing and social distancing, aim to reduce disease spread in the workplace. Our framework is designed to be adaptable to a variety of specific workplace environments to support planning efforts as reopenings continue.

Using this model, we consider a number of case studies, including an office workplace, a factory floor, and a university campus. Analysis of these cases illustrates that continuous testing can help a workplace avoid an outbreak by reducing undetected infectiousness even in high-contact environments. We find that a university setting, where individuals spend more time on campus and have a higher contact load, requires more testing to remain safe, compared to a factory or office setting. Under the modeling assumptions, we find that maintaining a prevalence below 3% can be achieved in an office setting by testing its workforce every two weeks, whereas achieving this same goal for a university could require as much as fourfold more testing (i.e., testing the entire campus population twice a week). Our model also simulates the dynamics of reduced spread that result from the introduction of mitigation measures when test results reveal the early stages of a workplace outbreak. We use this to show that a vigilant university that has the ability to quickly react to outbreaks can be justified in implementing testing at the same rate as a lower-risk office workplace. Finally, we quantify the devastating impact that an outbreak in a small-town college could have on the surrounding community, which supports the notion that communities can be better protected by supporting their local places of business in preventing onsite spread of disease.

## Introduction

The COVID-19 pandemic is a global crisis, with a devastating impact on people, organizations, and industries across the world. Efforts to reignite economic activity require a robust and safe return-to-work strategy. The signs and symptoms that characterize the disease vary, the mechanics of immunity are not fully understood, and a vaccine is still not available in most parts of the world.^1^ Additionally, a large proportion of infected individuals may never experience symptoms and can silently spread the disease.^2^ Therefore, an approach based solely on symptom tracking and testing of symptomatic individuals will be insufficient to prevent spread in most circumstances. Instead, augmenting symptom-based testing with cost-effective monitoring testing of the workforce has been proposed as a more promising strategy.^2^

Ideally, an employer must consider several factors when selecting a testing strategy. The disease prevalence in the surrounding community, and the rate of change of this prevalence, will impact the prevalence among employees and should thus be accounted for. Furthermore, the choice of testing strategy should incorporate features of the workplace such as the degree of close-contact interactions between employees and the amount of time that employees spend at work. In addition to selecting testing strategies for symptomatic and asymptomatic individuals, employers must make choices about how many employees they will bring back to work, and they must also consider the requirements for employees who test positive, such as the amount of time that they are asked to self-isolate away from the workplace.

These numerous considerations highlight the need for models that enable employers to anticipate, explore, and decide on policies that are appropriate for the particulars of their workplace. Such models can present the projected impact of various testing strategies, allowing an employer to make an informed decision on the most appropriate strategy. Models that give these insights have been explored in a university setting^3-7^ and in a healthcare setting^8^ but have not been thoroughly explored across different workplace settings.

An important component of virus spread in a workplace is the level of interaction of employees with non-employees in the community. This consideration is also important in a university setting, although since college campuses are often relatively self-contained a model may choose to ignore the ongoing influence of the community. Lopman et al.^6^ and Lyng et al.^7^ capture the impact of the community by including a continuous rate of spontaneous infection in the university population. Paltiel et al.^4^ instead add regular exogenous ‘shocks’ of infection to the university population to simulate the impact of the community, while Gressman et al.^5^ include a 25% chance that one member of the university population becomes spontaneously infected each day. However, none of these approaches are able to capture the time-varying impact of a community that is undergoing an outbreak and are also not able to capture the impact of the workplace on the community.

We present here a novel compartmental epidemiological model that accounts for SARS-CoV-2 transmission both within the workplace and in the surrounding community. This model is intended for use in forecasting prevalence in a workplace and guiding its choice of testing strategy. This model is designed to simulate how testing can be used alongside education and other workplace nonpharmaceutical interventions (NPI), such as masking policies, increased spacing of desks, and staggered return-to-work schedules, to allow workplaces to resume on-site activities while minimizing the risk of a new outbreak. Note that, in this paper, we use the term “outbreak” to refer to out-of-control spread of the virus, rather than a specific number of infected cases. We apply this model to investigate disease dynamics upon reopening of various workplace and university environments, demonstrating the flexibility of our approach in understanding disease spread and devising testing plans.

## Methods

We leverage a dynamic, deterministic, two-group thirteen-compartment model (Figure 1), which contains a SEPAYR (Susceptible - Exposed - Presymptomatic - Asymptomatic - sYmptomatic - Recovered) model for non-employees (“community”), alongside a SEPAYDR (Susceptible - Exposed - Presymptomatic - Asymptomatic - sYmptomatic - Detected - Recovered) model for employees (“workplace”). This “Community-Workplace” model accounts for transmission dynamics within and between the workplace and the community, and it can be used to simulate disease dynamics and inform the selection of a testing strategy for a specific workplace. For full details on the model and its parameters, please refer to Supplement S1. For details on how model parameters are chosen, see Supplement S2.

**Figure 1:**
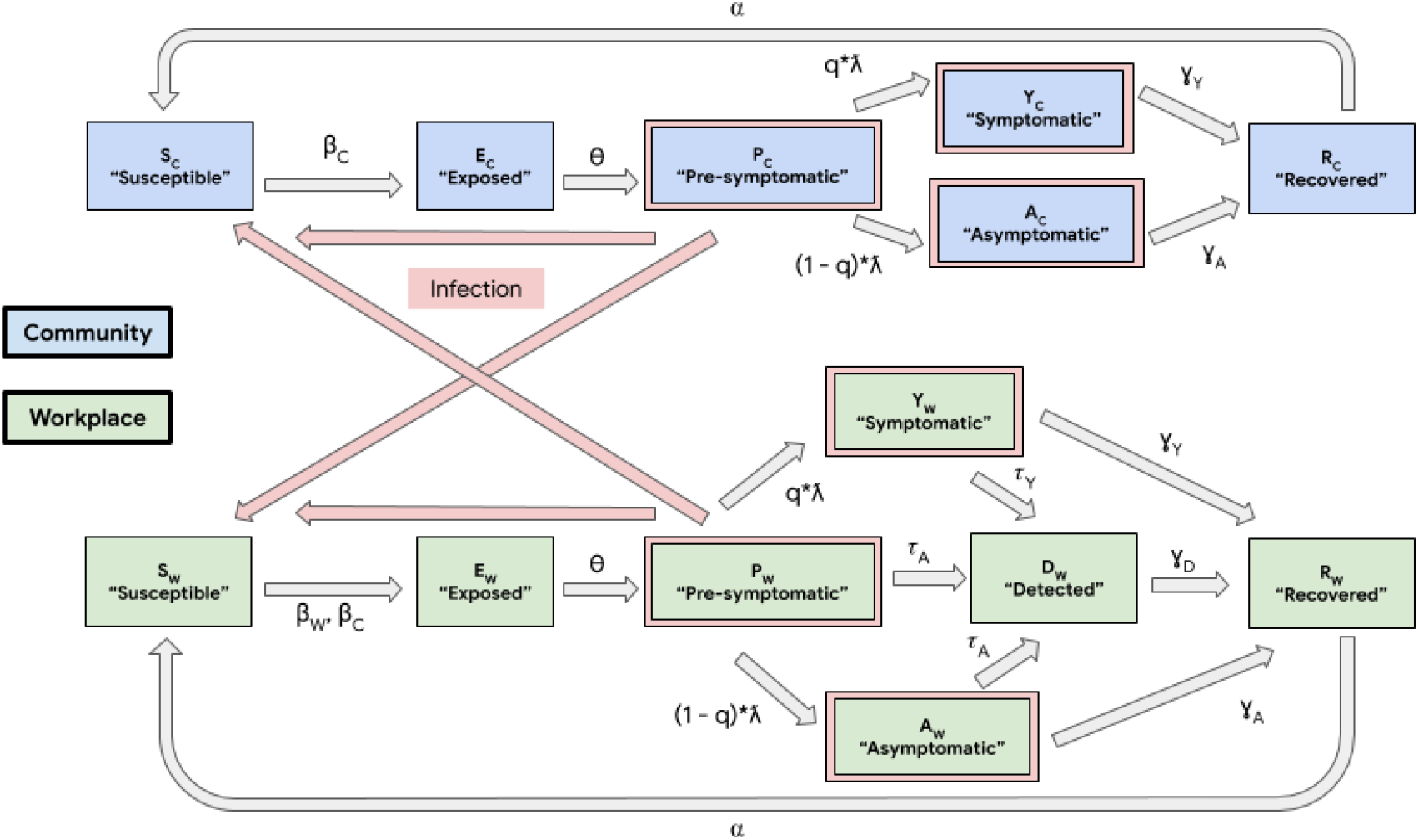
“Community-Workplace” compartmental model of disease spread in the workplace and community. The non-employees (“Community”, shaded blue, denoted with subscript “C”) are modeled using SEPAYR (Susceptible - Exposed - Presymptomatic - Asymptomatic - sYmptomatic - Recovered) compartments. The employees (“Workplace”, shaded green, denoted with subscript “W”) additionally can move through a Detected compartment, resulting in a SEPAYDR (Susceptible - Exposed - Presymptomatic - Asymptomatic - sYmptomatic - Detected - Recovered) model that tracks the stages of COVID-19 infection and detectability by workplace testing. Compartments of individuals that are sources of infections are outlined in pink, and pink arrows denote the paths of potential infections, i.e., disease transmissions. Model transition rate parameters are denoted on compartment-to-compartment transition arrows, and their semantics are detailed in Table S1.1.

To demonstrate the broad applicability of our modeling approach in the real world, we examined three case studies capturing some of the diversity of businesses and institutions of higher education in the United States:

a. Office workplace (representing a “9-to-5” workplace with lower density / contact load)
b. Factory floor (representing a “9-to-5” workplace with higher density / contact load)
c. University campus (representing an institution where many of the population spend a majority of their time, including sleeping, and where the population experiences higher density / contact load)

There are two key model parameters that are varied to emulate the environment for each case study. These are the basic virus reproduction number (i.e., the mean number of people in a fully susceptible population that are infected with SARS-CoV-2 by a single infected person) in the workplace (R0_W_), and the proportion of time employees spend at work and interacting only among themselves (p). Note that all parameters not used to capture this variation across the different environments were held constant across all case studies herein (see Table S1.2), except where noted below.

An R0_W_ of 3 was used to simulate an indoor “Office workplace” with a medium burden of employee-employee interactions, along with a value of p of 33%. A higher R0_W_ of 4 was used to simulate a “Factory floor” to capture the higher interaction between employees, due in part to increased physical density. As in the “Office workplace”, a value of p of 33% was used. An R0_W_ of 4 was used to simulate a “University”, to capture the heightened level of interaction expected between students who are living and attending classes together, as well as socializing after school hours. To capture the higher amount of time that on-campus students spend in one another’s company, a value of p of 70% was used.

For each case study scenario, a range of testing strategies were simulated and compared. Modeling a variety of testing strategies assists in the decision-making process, by yielding insights into the potential impact of the different strategies.^9^ The strategies investigated in our case studies, ordered by increasing testing volume, are as follows:

- **NO TESTING**: No testing at all.
- **INITIAL TESTING ONLY (I)**: Initial testing only (“back to work testing”).
- **I + SYMPTOMATIC TESTING (S)**: Initial testing, along with testing any employee that develops and self-reports symptoms, i.e., all symptomatics (see Supplement S2 for details on the choice of parameter defining the proportion of cases that develop symptoms).
- **I + S + TEST 5% OF ASYMPTOMATICS EVERY WORK DAY**: Initial testing, testing of all symptomatics, and testing of a randomly selected 5% of asymptomatic individuals each “work day” (i.e., 5 days a week). This strategy results in all asymptomatics being tested approximately <u>once every four weeks</u>.
- **I + S + TEST 10% OF ASYMPTOMATICS EVERY WORK DAY**: Initial testing, testing of all symptomatics, and testing of a randomly selected 10% group of asymptomatic individuals each work day (i.e., 5 days a week). This strategy results in all asymptomatics being tested approximately <u>once every two weeks</u>.
- **I + S + TEST 20% OF ASYMPTOMATICS EVERY WORK DAY**: Initial testing, testing of all symptomatics, and testing of a randomly selected 20% of asymptomatic individuals. This strategy results in all asymptomatics being tested approximately <u>once every week</u>.
- **I + S + TEST 40% OF ASYMPTOMATICS EVERY WORK DAY**: Initial testing, testing of all symptomatics, and testing of a randomly selected group of 40% of asymptomatic individuals. This strategy results in all asymptomatics being tested approximately <u>twice per week</u>.

A key feature of epidemiological models such as the one described here is that each of the variables of interest, in particular the number of individuals in each compartment, is tracked throughout the simulation. This permits the user to calculate and monitor a variety of metrics that can assess the projected severity and impact of COVID-19 outbreaks. For simplicity, we focus here on the prevalence of cases of active infection among employees (Equation S1.15). This metric captures the simultaneous impact of infected employees who are detected by testing and must miss work, alongside infectious employees at work that form a pool of active risk of exposing other employees to SARS-CoV-2.

## Results

Figure 2 shows the time-based trajectories of infection prevalence in the workplace population for the corresponding case studies, generated using the “Community-Workplace” model.

**Figure 2:**
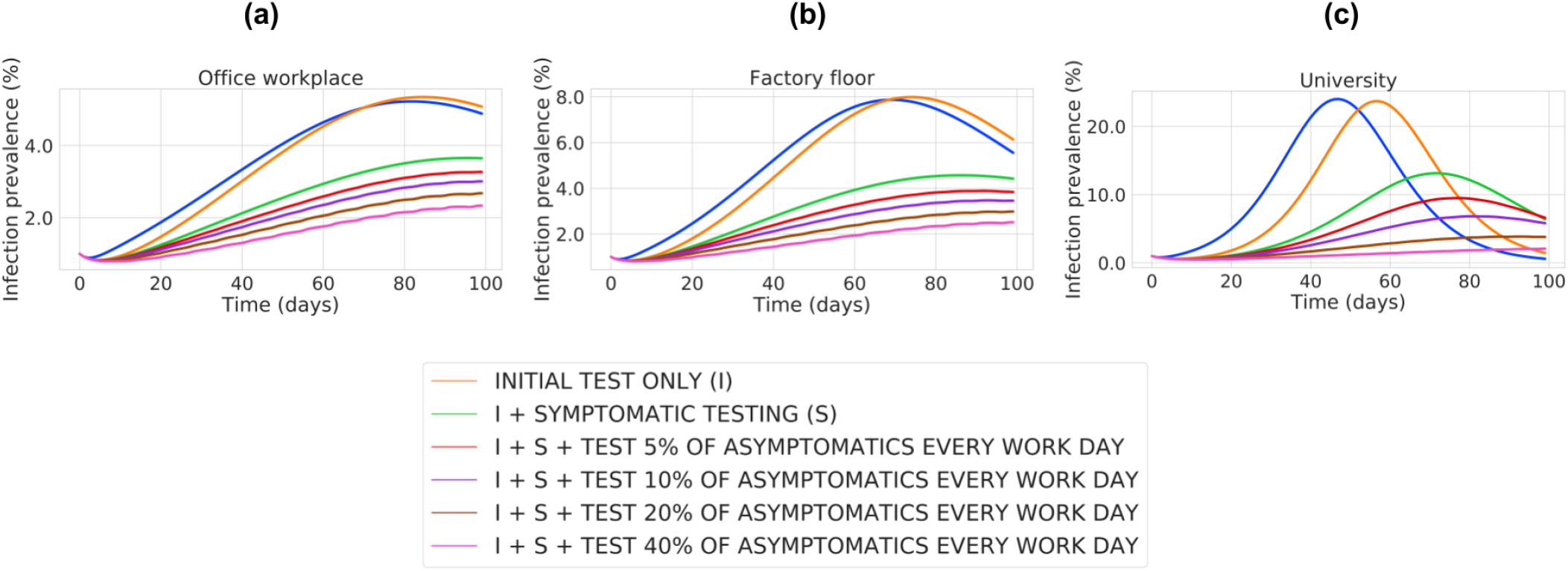
Estimated prevalence trajectory for the three case studies, each under a range of testing strategies. For each case study, the prevalence of employee infection is plotted on the vertical axis (as percentage of the total workplace population), tracked over time (in days) over the course of the simulation on the horizontal axis. For each scenario, the prevalence for the various assessed testing strategies are plotted as distinct curves and colored as per the legend. The parameters used for each case study are described in the main text, with more detail given in Supplement S1.

Other related metrics of interest are depicted in Supplement S3, including peak workplace prevalence (Figure S3.1), cumulative workplace prevalence (Figure S3.2), cumulative community prevalence (Figure S3.3), and total number of workplace tests conducted (Figure S3.4).

The “Office workplace” and “Factory floor” case studies (Figure 2a,b) differ only by the value of the parameter related to the transmission rate between employees in the workplace, R0_W_. Comparing these case studies, we see that increasing the transmission rate within the workplace (R0_W_ = 3 in the “Office workplace” vs. R0_W_ = 4 in the “Factory floor”) leads to higher peak prevalences (Figure S3.1a,b), as well as higher cumulative prevalences, i.e., total employees infected (Figure S3.2a,b). However, we find that increasing the testing volume diminishes these differences between workplaces. Specifically, when only symptomatic testing is conducted, the peak workplace prevalence in the two scenarios varies by 0.9%. When monitoring testing of 10% of asymptomatic people per day is added, the peak workplace prevalence in the two scenarios varies by 0.5%, and this difference drops to only 0.2% when 40% of asymptomatic people are tested each day.

The model includes a parameter, p, that describes the proportion of time that employees spend at work (see Table S1.1). By defining the amount of time that employees spend interacting only with one another, this parameter modulates the amount of infection spread between the workplace and community populations. To understand the importance of this parameter, we compare the “Factory floor” and “University” case studies (Figure 2b,c), since all other parameters are held constant. Spending a higher proportion of time isolated in a high-contact workplace environment (in the “University”) increases peak and total infections in the workplace/campus population (Figure S3.1b,c), and the “University” requires significantly more testing to achieve parity with the “Factory Floor”. Specifically, when testing 5% of asymptomatic individuals each work day, a peak prevalence of 3.9% is achieved in the “Factory Floor” setting, but 20% of asymptomatic individuals must be tested each work day in order to match this in the “University” setting. Of note, due to diminished interaction between the workplace and the community, the total percentage of individuals infected in the community is actually lower in the “University” case compared to the “Factory floor” case (Figure S3.3b,c).

A simultaneous comparison of these three case studies is instructive with respect to the role of testing. It demonstrates that as more time is spent together in a high-contact workplace environment, more aggressive testing of asymptomatic individuals is required to keep infection at safe levels. As an example, consider a 3% peak prevalence in the workforce as a high but still tolerable threshold. To maintain prevalence below that level, the “Office workplace” must test 10% of asymptomatic individuals per work day, the “Factory floor” must test 20% of asymptomatic individuals per work day, and the “University” requires testing of as many as 40% of asymptomatic individuals per work day (Figure S3.1).

A practical benefit of continuous testing strategies is that test results can be aggregated to derive an ongoing measure of prevalence in the workplace. If the employer closely follows such metrics, then mitigation strategies, such as augmentation of personal protective equipment and other NPI, can be introduced in a timely manner. To understand the impact of such interventions, we use our model to study how mitigations can impact the time dynamics of disease prevalence when they are introduced at a predetermined level of an “outbreak”. In Figure 3, we show how the trajectory of disease spread can be altered for the “University” setting with a constant testing strategy (everyone tested approximately every 2 weeks), but where additional mitigations are initiated as a response to the prevalence reaching an “unacceptably high” level. We find that introducing mitigations at 2% prevalence can reduce peak prevalence from 6.8% (Figure 3, purple curve) to 2.3% (Figure 3, gray curve). Of note, this is roughly equivalent to the case where mitigations are not introduced but instead testing is performed at fourfold the level, i.e., everyone is tested approximately twice per week (Figure 3, pink curve; see also Figure S3.1c).

**Figure 3:**
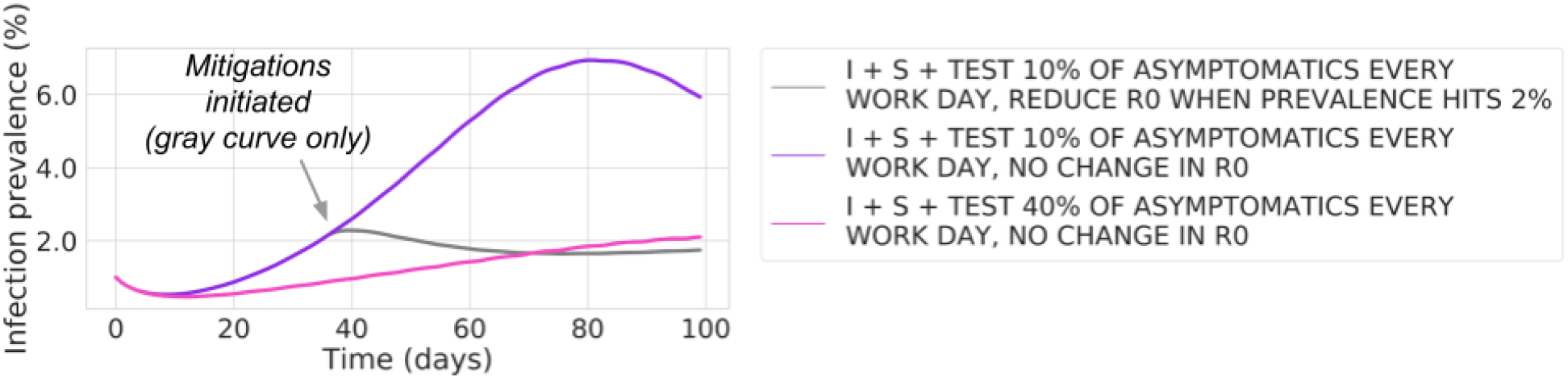
Workplace prevalence trajectories in the “University” case study, where workplace transmission can decrease as a result of mitigations introduced upon an increase in prevalence. Continuous testing allows an employer to estimate the current prevalence of infection in the workplace. This gives the employer the opportunity to introduce mitigations in response to increases in prevalence, reducing workplace spread. The gray curve shows the impact on workplace prevalence of testing 10% of asymptomatics per day but also reducing the workplace reproduction number (R0_W_, see Table S1.1) from 4 to 2 (corresponding to initiating mitigations) when prevalence reaches 2%; note that prevalence trajectories are plotted as in Figure 2. The purple curve (10% asymptomatic testing with no change in R0_W_ during the simulation) corresponds to the purple curve in Figure 2c, and the pink curve (40% asymptomatic testing with no change in R0_W_ during the simulation) corresponds to the pink curve in Figure 2c.

This analysis emphasizes the two distinct benefits of continuous testing: (i) detection and isolation of infectious individuals, directly suppressing disease spread; (ii) use of aggregated test results to estimate infection prevalence in the workplace, allowing an outbreak to be recognized in its early stages so that mitigations can be rapidly deployed. As noted above, an “Office workplace” can maintain prevalence below 3% by testing its workforce approximately every 2 weeks, whereas a “University” without dynamic mitigations requires its population to be tested as much as fourfold as often to achieve that goal (pink curves in Figures 2c,3).

However, as shown in Figure 3, a “University” that performs ongoing monitoring testing and can quickly react to a growing outbreak at 2% can maintain a prevalence below 3% (Figure 3, gray curve), yet requiring only as much testing as an “Office workplace” that has the advantage of lower contact load and not having people living together full time.

The “University” setting modeled in the above simulations assumes a community population of 500,000. Thus, it is more relevant to a university campus in a medium to large city rather than to a small college town. To understand the impact of community size on transmission, we repeated the “University” simulation using a community population of only 3,000 people, while maintaining the campus (“workplace”) population at 1,000. For this analysis, we assumed initial testing before return to campus, as well as ongoing testing of symptomatic individuals (but no randomized testing of asymptomatics). As shown in Figure 4a, the smaller community size does not have much impact on transmission on campus. However, it does cause a large increase in peak prevalence in the community (Figure 4b), from 2.8% with a community population of 500,000 to 12.8% for the community population of 3,000.

**Figure 4:**
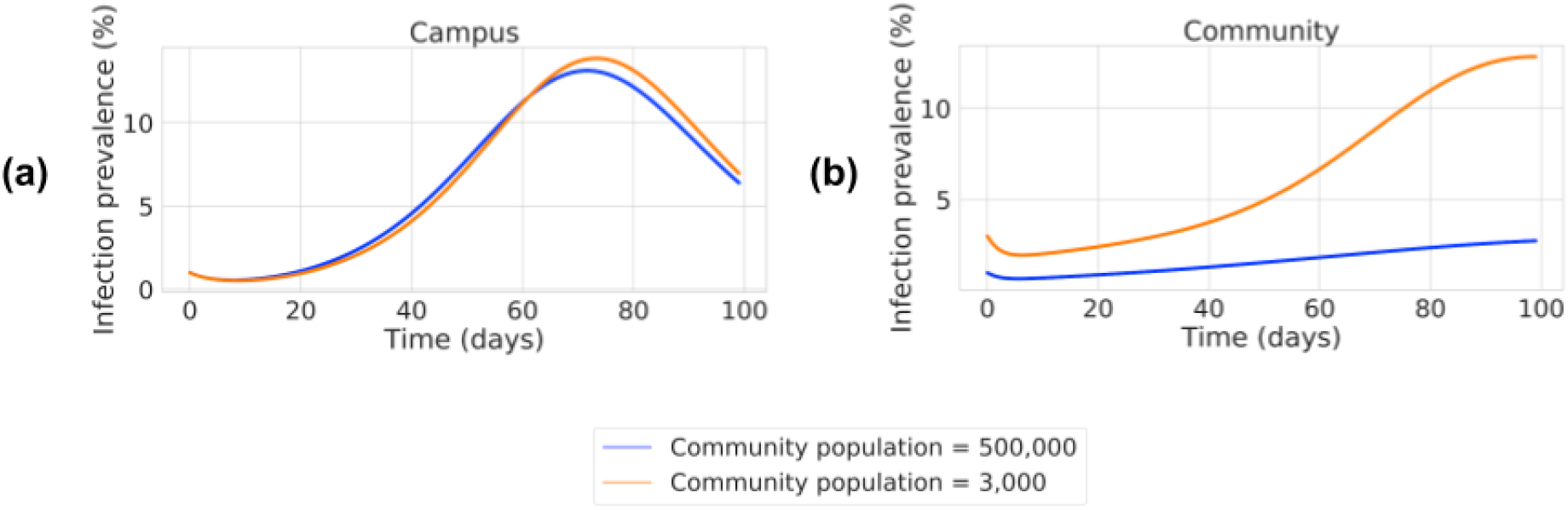
Impact of varying community population size on the workplace and community prevalence trajectories in a “University” setting with no asymptomatic testing. Prevalence trajectories (plotted as in Figure 2) for campus (“workplace”, panel a) and community (panel b) populations, for scenarios where initial testing and symptomatic testing are performed in the “University” (I + SYMPTOMATIC TESTING). The size of the community population here is either 500,000 (as used in all other simulations, blue curves) or 3,000 (orange curves). The campus (“workplace”) population is held constant at 1,000, as are all other simulation parameters as per Table S1.2. Thus, the blue curve in panel (a) simulates the same scenario of campus prevalence for a “University” in a larger city, as in the green curve in Figure 2c.

An advantage of analyzing results from the simulations performed herein is that we can easily tally the sources of infections, in contrast to real-world infections where it is quite difficult to make such determinations. For the community population of 500,000, a total of 91,834 community members (18.4% of the community [Figure S3.3c]) are infected during the simulation. We find that only 0.16% of these community infections are due to direct infection from the university population. In marked contrast, with the small community population of 3,000, a total of 719 community members (24.0%) are infected over the course of the simulation. Here, we find that 15.2% of these infections are directly attributable to disease transmission from the university. That is, these transmissions into the community arise from interactions between the infected university population and the susceptible community population, followed by community spread that ultimately results in a markedly increased community prevalence. Overall, these results illustrate the advantage of explicitly capturing the time dynamics of interactions between the community and the workplace.

## Discussion

Effective testing strategies are critical in allowing workplaces and schools of higher education to resume selected on-site activities, while minimizing the risk of outbreaks. The compartmental model presented here can be used to project how various testing strategies may impact the prevalence of infection in the workplace over time, allowing workplaces and schools to make more informed decisions about which testing strategy is best for them. Moreover, the model yields insights regarding when to introduce NPI mitigations when an outbreak is detected.

The compartmental model presented in this paper contains a SEPAYR model to track the epidemic in the community and includes interactions between this community population and the workplace population. In contrast, recent studies with similar goals have used single or continuous infection “seeds” to model infection originating from the community.^3,4,8^ While such a simplification allows the impact of varying workplace parameters to be studied, it does not capture the impact of changes in community prevalence on the workplace. Modeling the community also allows the model to capture the impact of workplace spread on the surrounding community. This becomes particularly important in a setting such as a small “college town” (Figure 4), where the size of the “workplace” population (i.e., students and university staff) is of a similar magnitude to the size of the surrounding community.^10^ More generally, because the safety of a community is dependent on lack of spread within its local workplaces, these results support the argument that it is in the interest of communities to support their places of business in preventing onsite spread of disease.

While the model presented in this paper gives a simplified view of the dynamics of COVID-19 infectiousness and transmission, the general framework provides a foundation for addressing real-world community-workplace scenarios. However, we emphasize that the assumptions inherent in this modeling approach must be clearly described to those who use its output to guide decisions. In Supplement S2, we review key considerations in understanding these parameters and some potential impacts of the values that are chosen.

Chief among such assumptions are the parameter choices that tailor the model to specific workplace settings. However, estimating the reproduction number (R0_W_, see Table S1.1) that will be experienced in a particular workplace setting is still an active area of research (see Supplement S2). Specifically, owing to the novelty of this virus, there is still only limited quantitative data on how and to what degree transmission occurs (e.g., by aerosol, contaminated surfaces, etc.^11^), especially in specific environments (e.g., in an air-conditioned office without fresh air but with social distancing and mask wearing).

The uncertainty that exists regarding the degree of disease transmission needs to be considered in the selection of an appropriate testing strategy. In particular, in comparing the “Office workplace” and “Factory floor” simulations, we found that more aggressive testing strategies control against outbreaks even in scenarios where there is higher contact in the workplace and thus diminish differences between higher and lower contact settings.

Therefore, when keeping prevalence extremely low is critical to ensure employee health and continued business operations, choosing as aggressive a testing strategy as possible (under budgetary and logistic constraints) will give the business the best chance of continuing to operate even with worst-case transmission rates.

In this work, we are somewhat conservative in the outcomes of the modeling (i.e., may overestimate infections) by not explicitly accounting for testing of community members, or for any degree of self-isolation undertaken by community members who exhibit symptoms. Nevertheless, such factors are implicitly accounted for in the choice of community and workplace R0 values.

For simplicity, the compartmental model we present here assumes a homogenous workforce population. However, in many workplace settings, there are distinct subgroups of employees with varied behaviors that result in differential infection risk. For example, such subgroups may include employees who work in different locations, employees who are customer-facing vs. those who have very little interaction with others, or university faculty vs. students. Employers may choose different testing strategies for these subgroups. Thus, a natural extension of the modeling here would be to permit such models to capture heterogeneity among the employees. This would likely require the addition of a set of new compartments for each subgroup, as well as parameters describing the rates of infection between every pair of subgroups. These complexities may make agent-based modeling^12^ better suited for this generalization. Similarly, generalizing to a heterogeneous community population would require the addition of analogous structure to the model. On a related point, age and other comorbidities have been shown to result in clinical heterogeneity once a person is infected;^13^ this model could be extended to account for clinical outcomes such as hospitalizations or deaths.

One of the purposes of testing (both symptomatic and monitoring of asymptomatics) is to detect infected individuals and remove them from the workplace in order to prevent workplace-acquired infections. As described above, another critical benefit of testing is to leverage the aggregated test results to continuously estimate infection prevalence in the workforce. By assessing workplace safety in real time, actions can be taken to prevent emerging outbreaks from growing (Figure 3). Such actions may include changes to existing mitigation measures (such as enforcing that personal protective equipment is properly used), or even shutting down the workplace if prevalence exceeds unacceptable thresholds. To ensure that such actions are taken in a timely manner, but only if necessary, it is critical that workplace prevalence be watched closely. In practice, the results of monitoring testing need to be translated to an estimate of workplace prevalence, with the statistical uncertainty around this estimate decreasing with larger sample sizes (i.e., with a larger volume of monitoring testing). Therefore, a return-to-work strategy that relies on quickly responding to nascent outbreaks benefits from a higher volume testing strategy that provides tighter statistical estimates of workplace prevalence. Such estimates would then be used to assess the safety of the workplace remaining open, for example, through either a formal statistical hypothesis test or by calculating a statistical distribution of likely prevalence values.

When deploying such testing programs in the real world, a key requirement for the employer is budgeting for the cost of the program despite the many uncertainties that the future holds. In our simulations, while the total number of tests performed varied substantially across testing strategies, the numbers for a particular strategy remained relatively stable across case studies (Figure S3.4). The main variation in the number of tests arises from testing of variable numbers of symptomatic individuals when outbreaks do occur. This relative stability of testing volume, for a fixed testing regimen, enables testing budgets to be estimated to a high degree of accuracy even before reopening a workplace, when parameters such as the reproduction number are still not known.

Repeated assessment of model adequacy is expected to be necessary due to the relatively short time horizon for which predictions can be reliably trusted due to the ever-changing state of societal policies and behaviors, as well as evolving clinical knowledge of the disease. Model parameters should be continuously updated to reflect scientific understanding of the disease, and also to reflect the observed test results and symptom reporting from the workplace of interest. This will allow the model output to be used to provide ongoing guidance about the projected impact of different workplace testing and other mitigation strategies. A concrete example of implementing such updates is the real-time estimation of the community virus reproduction number, R0_C_ (see Supplement S2).

## Conclusion

We present an epidemiological compartmental model that demonstrates the impact of testing strategies and dynamically-introduced employer mitigations on the spread of COVID-19 in a workplace. This model captures interactions between the workplace and the surrounding community population and can be tailored to fit the specifics of a wide range of workplace scenarios. To illustrate this flexibility, we present three case studies, simulating an office workplace, a factory floor, and a university campus. We discuss how to interpret insights from these simulations and how this model can guide the volume of testing intended to prevent workplace outbreaks from occurring or becoming large.

We also show how this modeling approach can allow employers to quantify how using ongoing testing can inform the real-time introduction of mitigations intended to prevent disease spread when outbreaks begin. In particular, we find that pairing data-driven mitigations with ongoing testing in a university can achieve the same benefit as substantially more testing. Additionally, we demonstrate how modeling the workplace and community populations together allows us to uncover important dependencies between these populations, which are particularly acute when the size of these populations is similar. In this setting, an outbreak in the workplace can lead to increased infection in the community, even when the community itself has mitigations in place to reduce transmission. Lastly, we reiterate that data from the workplace of interest should be used to adjust model parameters over time. This approach should improve model accuracy for continuous forecasting of disease prevalence and thus better empower employers to choose testing strategies that meet the goal of keeping their business up and running within explicit safety parameters.

## Data Availability

Any data sources referred to in the manuscript are external and publicly available and linked in the reference section.

## Supplementary Materials

### Supplement S1: Supplementary Methods

We present here a two-group compartmental model that accounts for transmission dynamics of SARS-CoV-2^1-3^ within and between two groups: (i) employees in the workplace (“the workplace”, denoted as “W”); (ii) non-employees (“the community”, denoted as “C”). The basic modeling framework rests on the following key assumptions (Figure S1):

a. **FIXED WORKFORCE**: All individuals are either employees or non-employees.
b. **VARIABLE SPREAD**: The transmission rate in the workplace and in the community may differ.
c. **COMMON BIOLOGY**: The progression of the disease, including time to develop symptoms and time to recover, is the same among employees and non-employees.
d. **SHARED COMMUNITY**: Employees spend p% of their time at work and isolated from the non-employees in the community. During the remaining (100 - p)% of time, employees interact with both employees and non-employees in the community (though their interactions may quantitatively differ).

**Figure S1:**
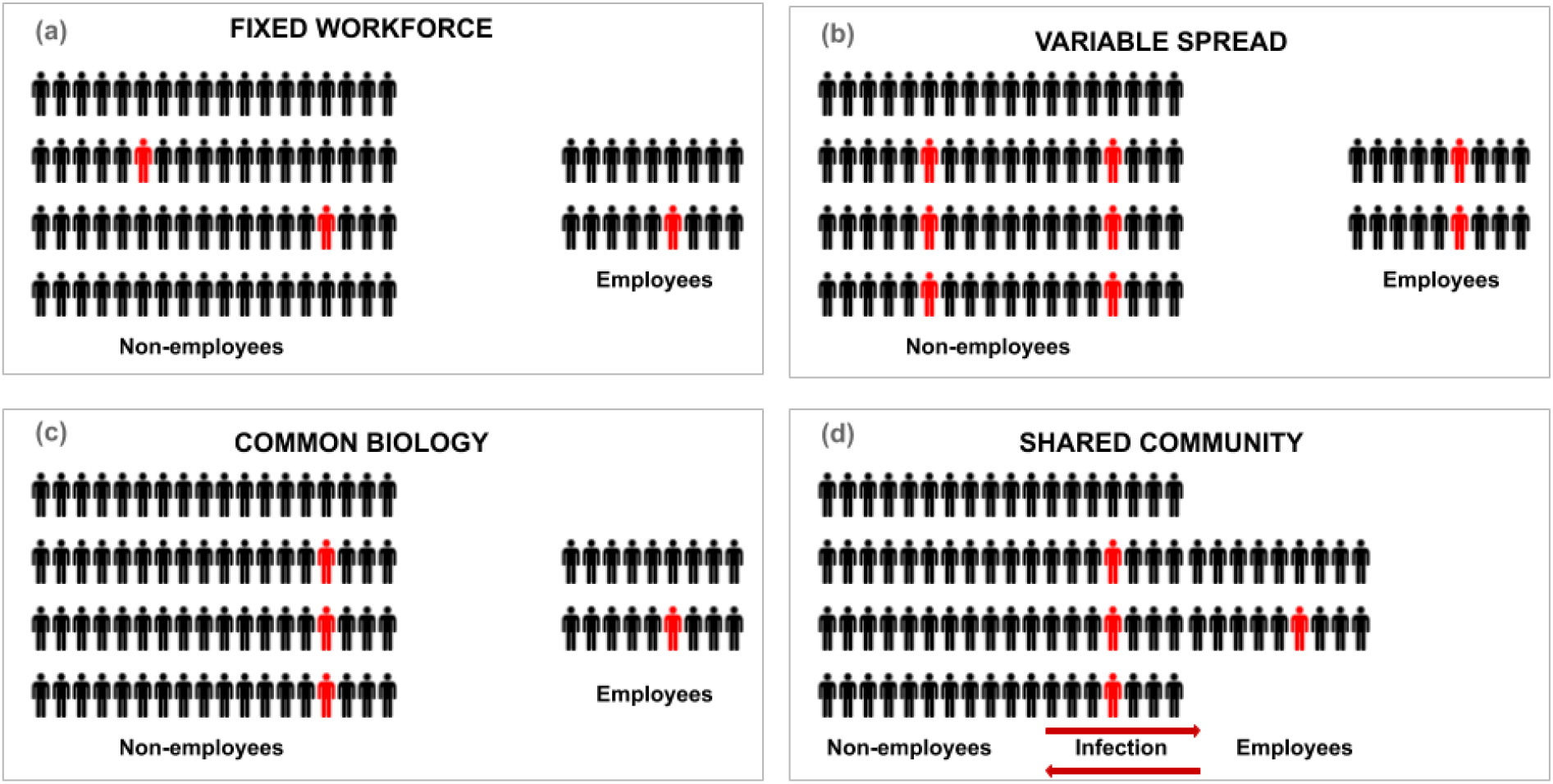
Foundational principles for modeling disease transmission dynamics within and between the community and the workplace. (a) FIXED WORKFORCE: People in the workforce remain as “employees” during the entire simulation, and all other individuals are considered “non-employees”. **(b) VARIABLE SPREAD**: Transmission dynamics among employees may differ from transmission among non-employees. In the cartoon example shown here, taking panel (a) as the starting point, the 2 infected out of 72 non-employees have infected twice as many non-employees (4), whereas the 1 infected of 18 employees has infected only 1 additional employee (perhaps due to stricter social distancing in the workplace). **(c) COMMON BIOLOGY**: In this example, taking panel (b) as the starting point, a week after their infections have started, half of the infected individuals have recovered, irrespective of being an employee or not. **(d) SHARED COMMUNITY**: Outside of the workplace, employees and non-employees mix and allow for transmission of virus within this larger group.

These assumptions are translated into a dynamic, deterministic, two-group compartmental model that we call the “Community-Workplace” model. This model is composed of a SEPAYR (Susceptible - Exposed - Presymptomatic - Asymptomatic - sYmptomatic - Recovered) compartmental model for non-employees and a SEPAYDR (Susceptible - Exposed - Presymptomatic - Asymptomatic - sYmptomatic - Detected - Recovered) compartmental model for employees (Figure 1, Table S1.1, and Equations S1.1 - S1.16).

**Table S1.1.** Notation and formulae for parameters used in the “Community-Workplace” model.

| Model Concept | Notation |
| --- | --- |
| Length of modeling simulation (days) | $L$ |
| Time (timepoint of the simulation) | $t$ |
| “Workplace” population size (i.e., number of employees) | $N_W$ |
| “Community” population size (i.e., number of non-employees) | $N_C$ |
| Initial disease prevalence in the community (at the beginning of the simulation) | $prv_{Ci}$ |
| Initial disease prevalence in the workplace (at the beginning of the simulation) | $prv_{Wi}$ |
| Proportion of time employees spend at work (interacting only among themselves) | $p$ |
| Proportion of cases that develop symptoms (vs. being “asymptomatic”) | $q$ |
| Proportion of non-cases that report symptoms each work-day | $g$ |
| Average days it takes to develop infectiousness after infection-causing exposure to virus | $\Delta_{infectious}$ |
| Rate of development of infectiousness ($= 1 / \Delta_{infectious}$) | $\theta$ |
| Average days of being infectious. Equivalent to the average days taken to recover from onset of infectiousness. | $\Delta_{recover}$ |
| Rate of recovery from onset of infectiousness ($= 1 / \Delta_{recover}$) | $\gamma_I$ |
| Average days taken to develop symptoms after becoming infectious (for non-“asymptomatic”s) | $\Delta_{symptoms}$ |
| Rate of symptom development for infected individuals who become symptomatic ($= 1 / \Delta_{symptoms}$) | $\lambda$ |
| Rate of recovery from onset of symptoms ($= 1 / (\Delta_{recover} - \Delta_{symptoms})$) | $\gamma_Y$ |
| Rate of recovery from moving into asymptomatic compartment ($= 1 / (\Delta_{recover} - \Delta_{symptoms})$) | $\gamma_A$ |
| Days required in isolation if tested positive | $\Delta_{isolation}$ |
| Rate of movement back to work after being detected ($= 1 / \Delta_{isolation}$) | $\gamma_D$ |
| Average days of immunity after recovering | $\Delta_{immunity}$ |
| Rate of loss of immunity ($= 1 / \Delta_{immunity}$) | $\alpha$ |
| Basic virus reproduction number (i.e., the mean number of people in a fully susceptible population that are infected with SARS-CoV-2 by a single infected person) in the workplace | $RO_W$ |
| Basic virus reproduction number (i.e., the mean number of people in a fully susceptible population that are infected with SARS-CoV-2 by a single infected person) in the community | $RO_C$ |
| Transmission rate in the workplace ($= RO_W * \gamma_I$) | $\beta_W$ |
| Transmission rate in the community ($= RO_C * \gamma_I$) | $\beta_C$ |
| Proportion of the asymptomatic workforce population tested each day | $\tau_A$ |
| Proportion of the symptomatic workforce population tested each day | $\tau_Y$ |
| Test sensitivity (probability of an infected individual's test being positive) | sens |
| Average number of tests required to return to work after infection (i.e., "testing out of isolation") | h |

Under this idealized model, susceptible individuals (S_C_ and S_W_, for non-employees and employees, respectively) can be infected by either employees or non-employees (P_C_, A_C_, and Y_C_, or P_W_, A_W_, and Y_W_, respectively; see below). This is achieved using a variable p that defines the amount of time the employees spend isolated among themselves in the workplace. During this time, employees only interact with one another, and as such, infection can only be transmitted from employee to employee. Similarly, non-employees only interact with one another during this time. In the remaining time (1 - p), the employees and non-employees act as one population, allowing infection to be transmitted between employees and non-employees. Upon becoming infected, individuals move into an exposed state (E_C_ and E_W_, for non-employees and employees, respectively). While in the exposed state, the viral load is considered too low to be detectable, and the individual is modeled as not yet being infectious. Once the viral load increases sufficiently, the individual becomes infectious and would also return a positive result from a diagnostic test that has 100% sensitivity. This is a simplification we use here of the relationship between viral load and the probability of testing positive, though there is reason to believe that test sensitivity increases as viral load increases, rather than having a distinct threshold.^4^ Infectious individuals fall into three compartments, presymptomatic (P_C_ or P_W_), asymptomatic (A_C_ or A_W_), and symptomatic (Y_C_ or Y_W_). Infected employees who test positive in the workplace testing program move into a detected compartment (D_w_). All infected individuals move to a recovered compartment (R_C_ or R_W_, for non-employees and employees, respectively) once viral load again drops to a non-infectious level; as noted above, for simplicity, this is equated with being below detectable levels for testing. This model assumes that symptomatic and asymptomatic individuals are equally infectious: for more detail on this assumption, see Supplement S2 below.

Using the notation defined in Table S1.1, the specified dynamics of this two-group “Community-Workplace” compartmental model are governed by the system of differential equations given in Equations S1.1 - S1.14. Note that the model used herein was implemented using discrete-time difference equations, but differential equations are shown here for clarity.

#### Non-employee SEPAYR

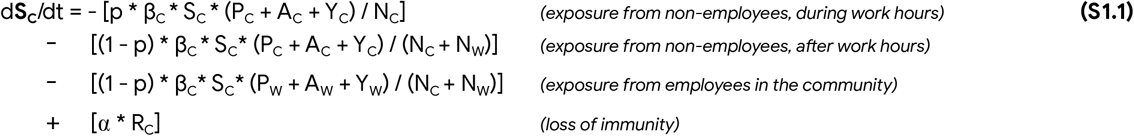

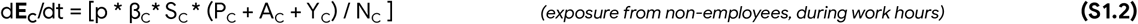

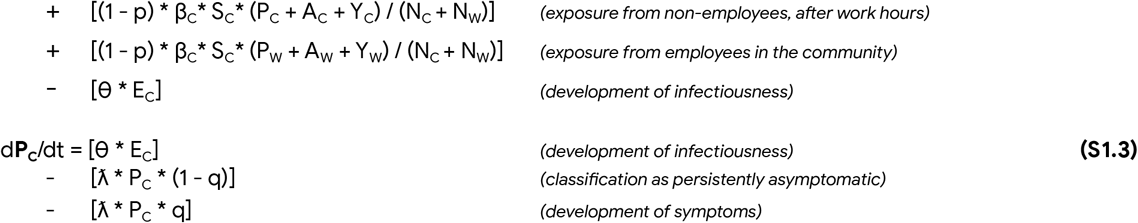

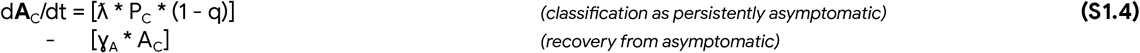

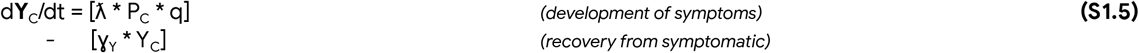

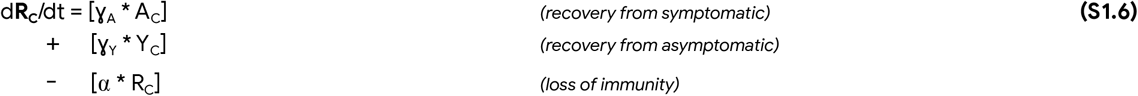

#### Employee SEPAYDR

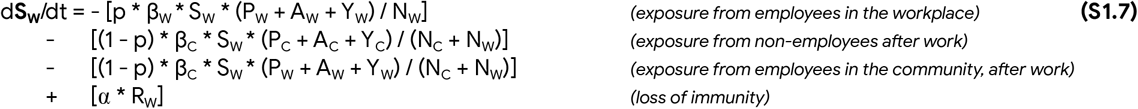

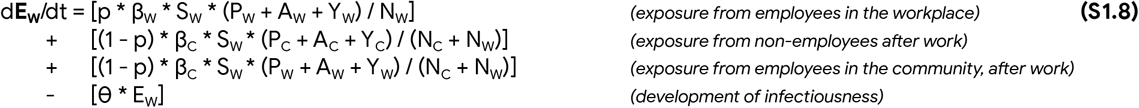

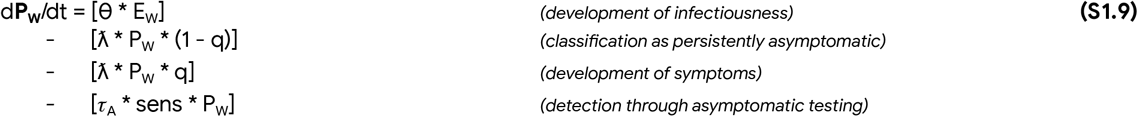

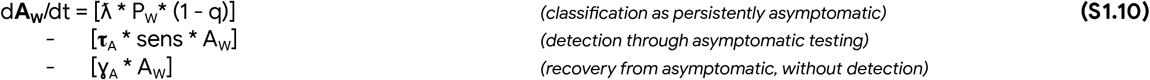

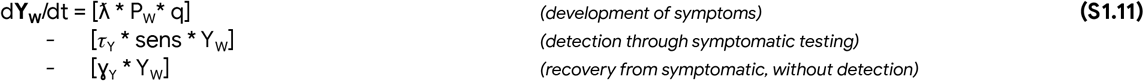

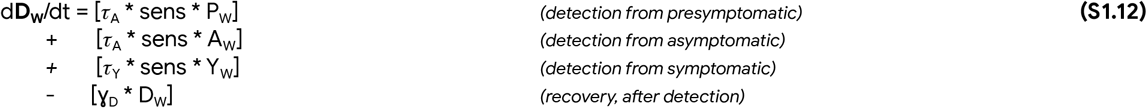

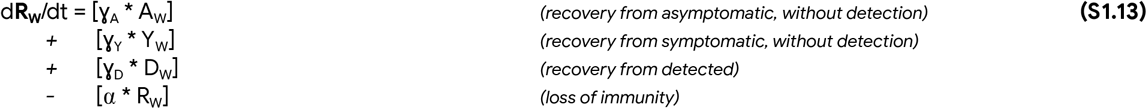

Of note, the efficacy of any real-world testing program in finding those who are infected is attenuated by the sensitivity of the test being used. Thus, all testing rates (τ) in Equations S1.9 - S1.12 are multiplied by the sensitivity of the test (“sens”), which is typically below 1.0.^5^

In parallel to the differential equations for compartmental transitions, our model also estimates the total number of tests that are performed over the time period of interest. The test count includes tests performed due to reported symptoms, random monitoring testing of the population, and recovery testing that allows isolated individuals to return to the workplace. Estimating the number of tests of symptomatic individuals requires making an assumption about the proportion of non-cases that report symptoms each day (g parameter in Table S1.1). Thus, this additional variable counting tests being performed is tracked as follows:

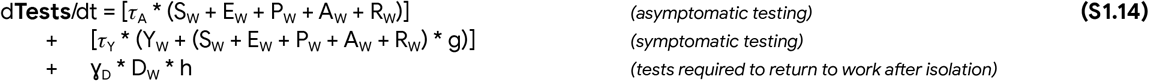

Note that Equation S1.14 includes individuals in the recovered compartment (R_W_) in both symptomatic and asymptomatic testing. The recovered compartment includes both individuals who moved from detected to recovered (“known recovereds”), and those who moved from symptomatic or asymptomatic to recovered (“unknown recovereds”). Because the unknown recovereds cannot be distinguished in practice from the susceptible population, an employer cannot actually ever choose to exclude all recovered individuals from testing. While we could add here an additional compartment to separate known and unknown recovereds (thus allowing known recovereds to be excluded from testing), this adds complexity to the model that is only necessary for the secondary analysis of counting tests. Instead, we have chosen for simplicity of exposition here to include all recovered individuals in ongoing testing.

Moreover, employers in practice may in fact choose such a conservative approach due to the unknowns regarding the degree of immunity conferred to individuals who have recovered from the virus, and also because there may be individuals who received a false positive result and so are incorrectly believed to be recovered from the virus.

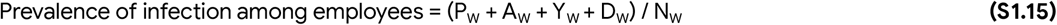

The main analyses presented in this paper focus on the prevalence of cases of active infection among employees (that is, those that have the potential to be infectious if they interact with other individuals). This quantity is calculated as:

Prevalence among non-employees is calculated analogously to Equation S1.15, though without the “Detected” compartment (since we are not modeling testing in the community).

For a given simulation run of the model, the initial state of the system is defined to match the current prevalence in the population of interest. To start, the size of the “Infected” compartment in the community sub-model is determined by the reported prevalence in the community. Next, recall that the model includes the option for the entire workforce to undergo initial testing prior to returning to the workplace. With a perfectly sensitive test, this approach would result in an initial employee prevalence of 0%. However, to account for imperfect test sensitivity,^19^ the expected initial employee prevalence can be calculated by multiplying the false negative rate of the test by the initial community prevalence:

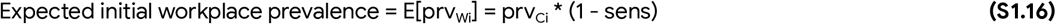

Equation S1.16 is used to initialize the workplace prevalence value when initial testing is selected as part of the overall testing strategy. On the other hand, if the option for initial testing prior to returning to work is not selected, the workplace prevalence is simply initialized to be equal to the starting community prevalence (prv_Ci_). In either case, the initial infected (but not detected) employee population is distributed between the “Symptomatic” and “Asymptomatic” compartments, according to the parameter q. Any infected individuals who were identified during an initial testing process begin the simulation in the “Detected” compartment. For simplicity, all other compartments are initialized to occupancies of 0.

In the Results section, we present 3 case studies that demonstrate the flexibility of the “Community-Workplace” modeling framework. These case studies involve configuring the model to emulate an office workplace, a factory floor, and a university. Table S1.2 gives the values of the model parameters that are held constant for all case studies, and in all follow-up simulations except where otherwise noted. In the main text, we described the choices of the R0_W_ and p parameters that are varied to capture an idealized portrayal of each case study. Thus, the parameters for each of the case studies are fully described by these case-specific values alongside the values in Table S1.2.

**Table S1.2.**
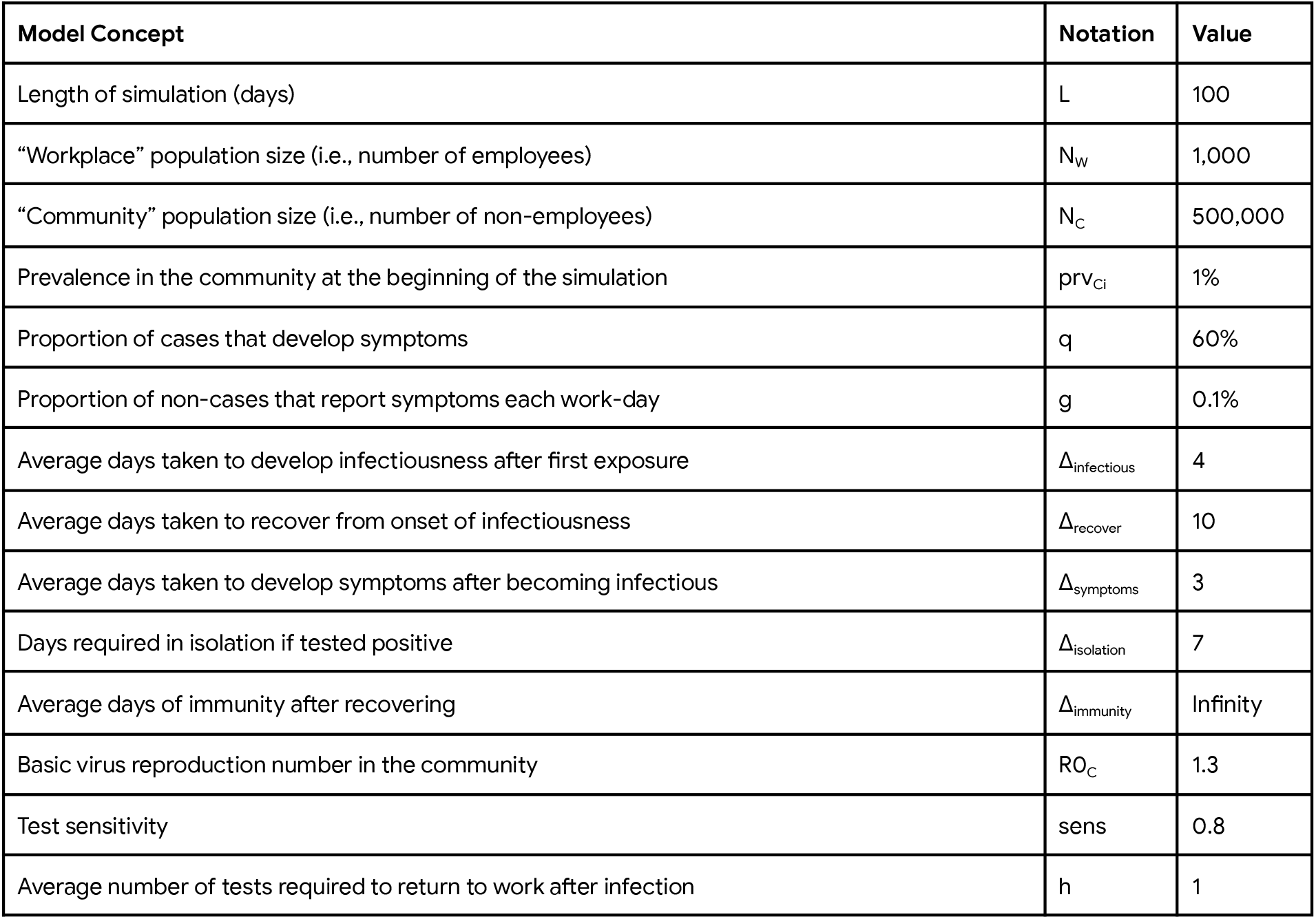
Model parameters held constant for all case studies. All model parameters not listed here or in the main text can be calculated from these values using the formulae in Table S1.1. See Supplement S2 for a discussion of how these values were selected.

### Supplement S2: Estimation of community and workplace model parameters

The compartmental model presented has a large number of parameters. Some of these parameters are related to epidemiological characteristics of the SARS-CoV-2 virus and the associated COVID-19 disease. Because this virus and the disease that it can cause are not yet fully understood, there is uncertainty in the values of these parameters. Other parameters are related to the initial prevalence and transmission rates in the populations of interest.

Translating a realistic scenario into the appropriate values of these parameters is challenging, as there does not yet exist enough data for us to accurately estimate how different workplace characteristics and policies (e.g., employee density, mask-wearing, and hand-washing) translate into epidemiological concepts such as R0. To ensure that the uncertainty in the value of the model parameters is reflected in the model output, we suggest performing a probabilistic sensitivity analysis, varying parameters over their expected range.^6^

The following sections specify how the default values of each model parameter are currently being estimated.

## Initial community prevalence

This parameter should reflect the total number of active cases in the community. Most US states and counties report the number of confirmed cases that are logged each day through publicly available channels. However, due to variation in testing availability and criteria in different geographic areas, it is not straightforward to translate from reported cases to active cases of infection.^7-9^ Therefore, this is still an area of active exploration in the research community. The value used for initial community prevalence in our case studies is based on an average value for the estimated prevalence in US states. These estimates were initially obtained from a publicly available model (covid19-projections.com) that fits an SIR-style model to reported case numbers and death rates to obtain estimated current prevalence for a geographic area of interest;^10^ note that this web server has stopped driving prevalence values and thus we used a conservative estimate of 1% for the generic examples considered here. We found that lowering this parameter in the community from 1% to, e.g., 0.5%, simply somewhat delayed the timing of the prevalence trajectories rather than qualitatively changing their properties, in particular since we use R0_C_ > 1.

### Community R0 (R0_C_)

Similarly, the estimate of the current R0 in specific geographic areas is best obtained using a model that incorporates multiple sources of data, including reported case numbers and death rates. The value of this parameter used in our case studies is obtained from the same model we used for community prevalence, and also from the Rt.live model.^10,11^

### Workplace R0 (R0_W_)

Estimating an appropriate value of R0 for a specific type of workplace is another area of active research. Currently, values of this parameter are informed by looking at existing studies of COVID-19^7,8^ and tracking of outbreaks within populations of interest.^13^

### Proportion of time spent at work

The proportion of time that employees spend at work determines the amount of spread between the community and the workplace. In workplaces where employees typically work a 40 hour week, we set this parameter to 33%. This assumes that the employee is spending two-thirds of their time as a member of the wider community. This includes time outside of work during the week, as well as time over the weekend. For the “University” case study, we consider that many students live in dorms on campus, attend classes together, and eat most of their meals in on-campus dining halls. As such, the default value for “time spent at work” in the “University” case study is set to 70%.

### The Time Course of Disease

Estimates of the time between exposure to the virus, development of infectiousness, and development of symptoms are based on CDC guidance, as well as values used in similar studies.^3, 12, 14^ We use slightly more conservative values, in that we assume in an average of 4 days between exposure and the virus becoming detectable (and infectiousness). Even if the full population is tested each day, infected individuals who are in this latent period will not be detected. Similarly, we assume an average of 3 days between the development of infectiousness and the development of symptoms. This is an increase on the estimate of 2 days seen in other studies,^3, 14^ meaning that infected individuals will remain in the workplace for a longer period of time. The choice of these parameters ensures that we err of the side of being slightly pessimistic about outcomes within a population.

### Proportion of cases with symptoms

The model parameter q indicates the proportion of cases that are expected to self-report symptoms. The current best estimate from the CDC^15^ is that 60% of COVID-19 cases will develop symptoms; we thus use this value.

### Lapsing of immunity (α)

The question of immunity after infection is still an area of research.^16^ Because the simulations conducted by our model are on a relatively short time scale (100 days), we currently assume that all recovered individuals maintain immunity to further infection for the duration of the simulation, so the parameter α is set to 0. Understanding the appropriate value for α will be critical to ensure that populations who have been infected and have recovered, as well as populations who have been vaccinated for COVID-19, can be effectively incorporated into the model.

### Infectiousness of asymptomatic individuals relative to symptomatic individuals

The model presented assumes that asymptomatic individuals are equally infectious as symptomatic individuals. However, the CDC believes there is some evidence that asymptomatic individuals are somewhat less infectious than symptomatics,^15^ and a difference in relative infectiousness is actually built into some published models^17^. While adding this extension to our model would not be technically difficult, we feel that assuming equal infectiousness is the correct choice for our use case. In particular, assuming equal infectiousness gives an conservative (pessimistic) view of the potential spread within a workplace if asymptomatic individuals are undetected, ensuring that the model does not give employers an overly optimistic picture of the likely trajectory of an epidemic within their workplace.

## Supplement S3: Case study results

**Figure S3.1.**
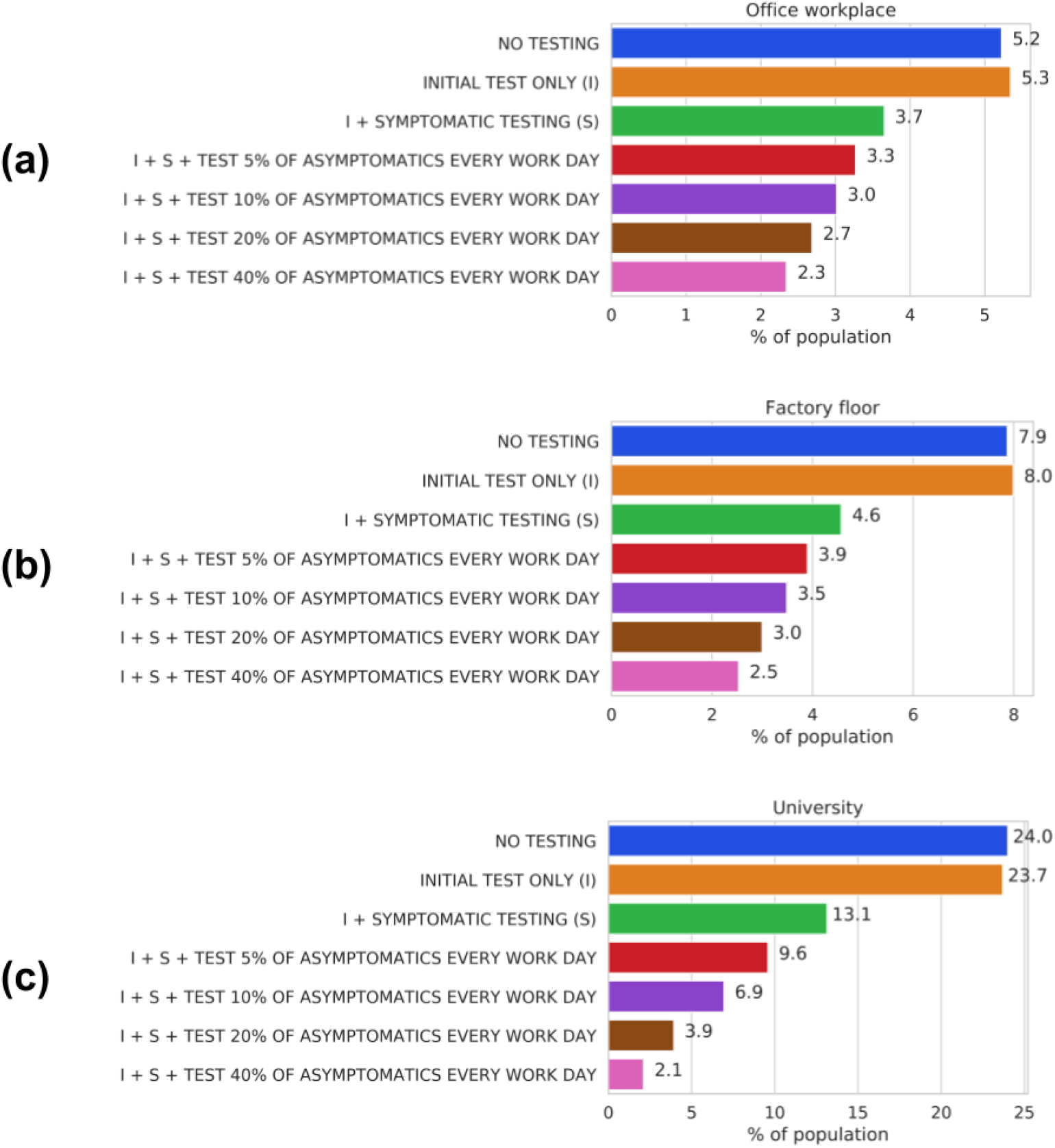
Peak workplace prevalence (as percentage) under a range of testing strategies, for (a) Office workplace, (b) Factory floor, (c) University.

**Figure S3.2.**
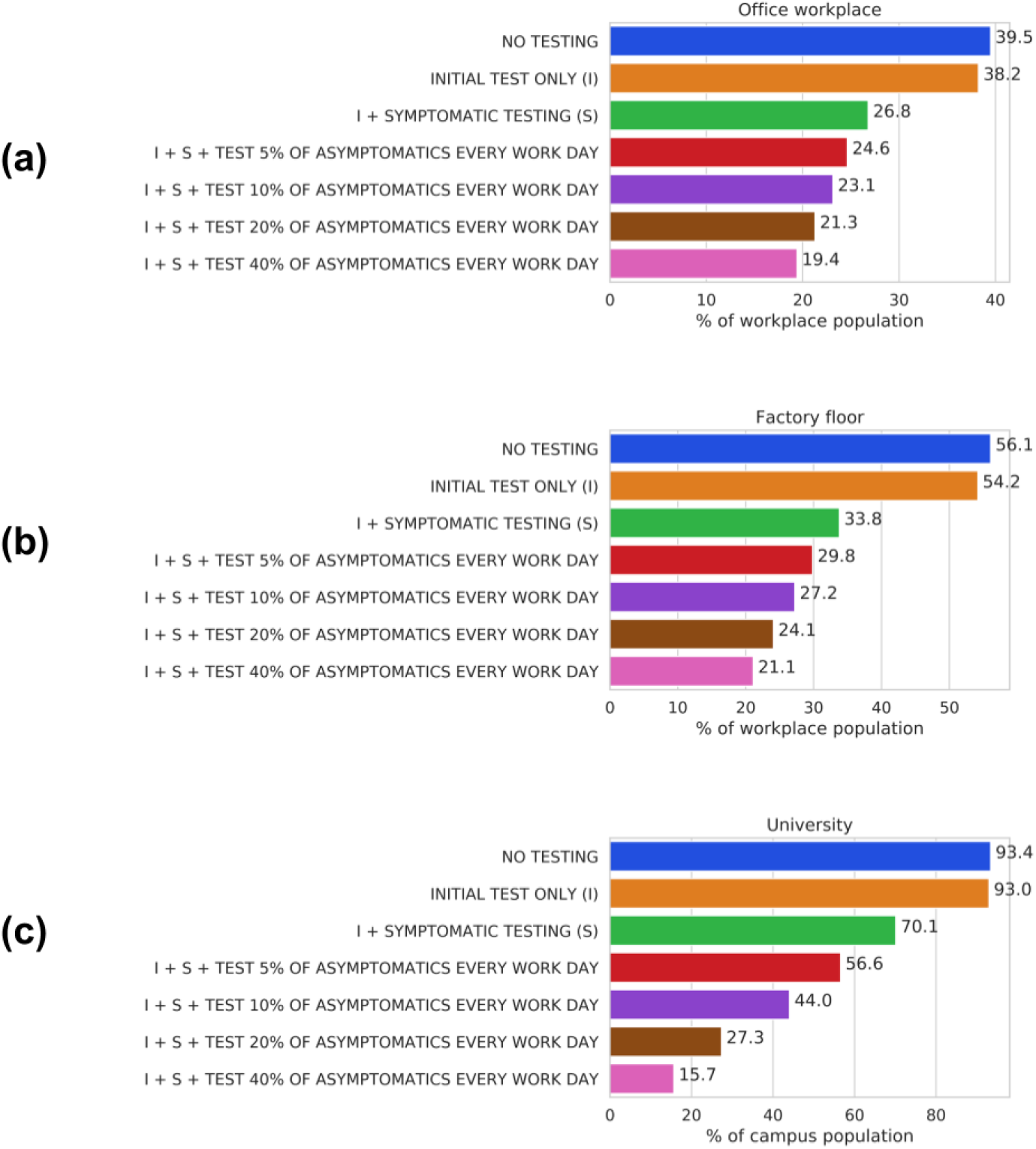
Percentage of workforce population infected over the entire simulation (cumulative prevalence) under a range of testing strategies, for (a) Office workplace, (b) Factory floor, (c) University.

**Figure S3.3.**
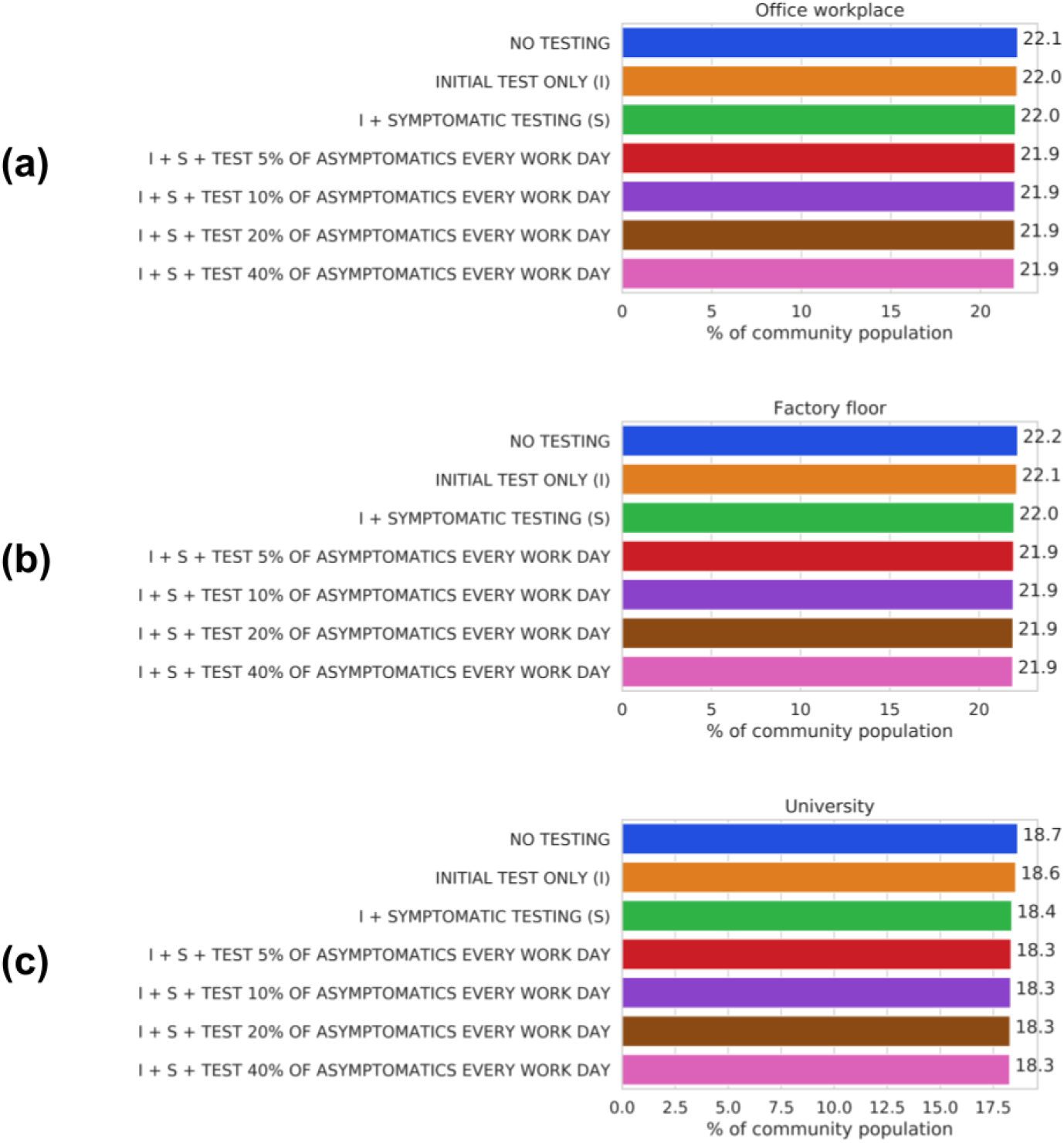
Percentage of community population infected over the entire simulation (cumulative prevalence) under a range of testing strategies, for (a) Office workplace, (b) Factory floor, (c) University.

**Figure S3.4.**
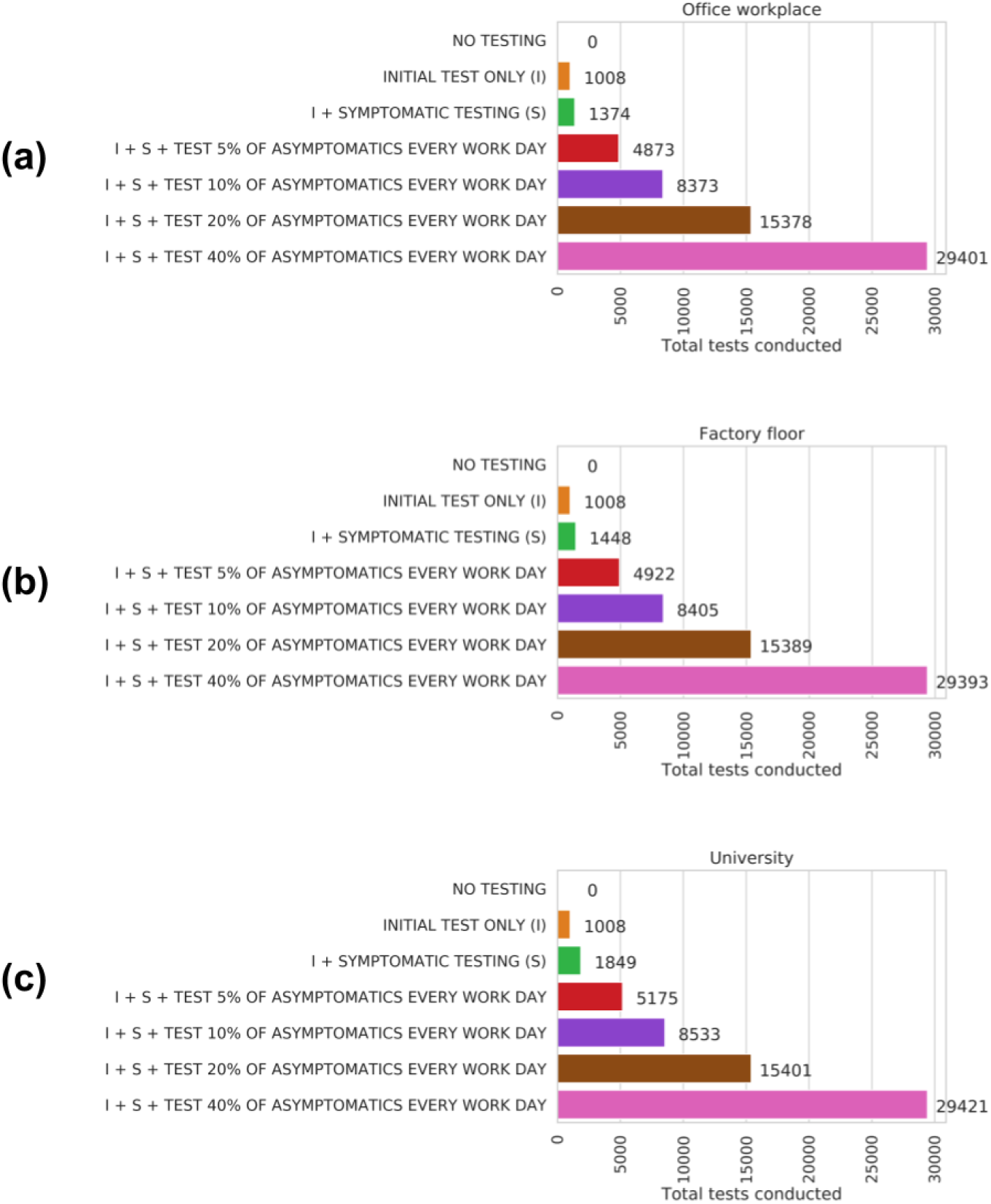
Estimated total workplace tests conducted over the 100 day simulation under a range of testing strategies, for (a) Office workplace, (b) Factory floor, (c) University.

